## Supplementary material for "Geographic differences in population rates of interventional treatment for prostate cancer in Australia": SuppInfo: Detailed description of the Methods and additional Results

#### Results

##### Statistical groups

*Table S1; Number and percentage of areas in each group by state, area-level socioeconomic quintile and remoteness category.*

| | High prostatectomy group | High EHR group | High LDR Brachytherapy group |
|---|---|---|---|
| Total | 875 (41%) | 739 (34%) | 534 (25%) |
| State or territory | | | |
| New South Wales | 378 (71%) | 143 (27%) | 13 (2%) |
| Victoria | 76 (18%) | 248 (58%) | 104 (24%) |
| Queensland | 304 (59%) | 175 (34%) | 38 (7%) |
| Western Australia | 0 (0%) | 50 (21%) | 187 (79%) |
| South Australia | 0 (0%) | 12 (7%) | 153 (93%) |
| Tasmania | 15 (16%) | 41 (43%) | 39 (41%) |
| Australian Capital Territory | 102 (97%) | 3 (3%) | 0 (0%) |
| Northern Territory | 0 (0%) | 67 (100%) | 0 (0%) |
| Area-level socioeconomic index | | | |
| Most disadvantaged | 119 (28%) | 220 (53%) | 80 (19%) |
| Q2 | 137 (33%) | 159 (38%) | 125 (30%) |
| Q3 | 151 (36%) | 168 (40%) | 100 (24%) |
| Q4 | 182 (43%) | 123 (29%) | 116 (28%) |
| Most advantaged | 267 (63%) | 54 (13%) | 100 (24%) |
| Remoteness category | | | |
| Major city | 589 (47%) | 354 (29%) | 299 (24%) |
| Inner regional | 211 (43%) | 163 (33%) | 115 (24%) |
| Outer regional | 67 (21%) | 152 (47%) | 102 (32%) |
| Remote and very remote | 8 (8%) | 70 (73%) | 18 (19%) |

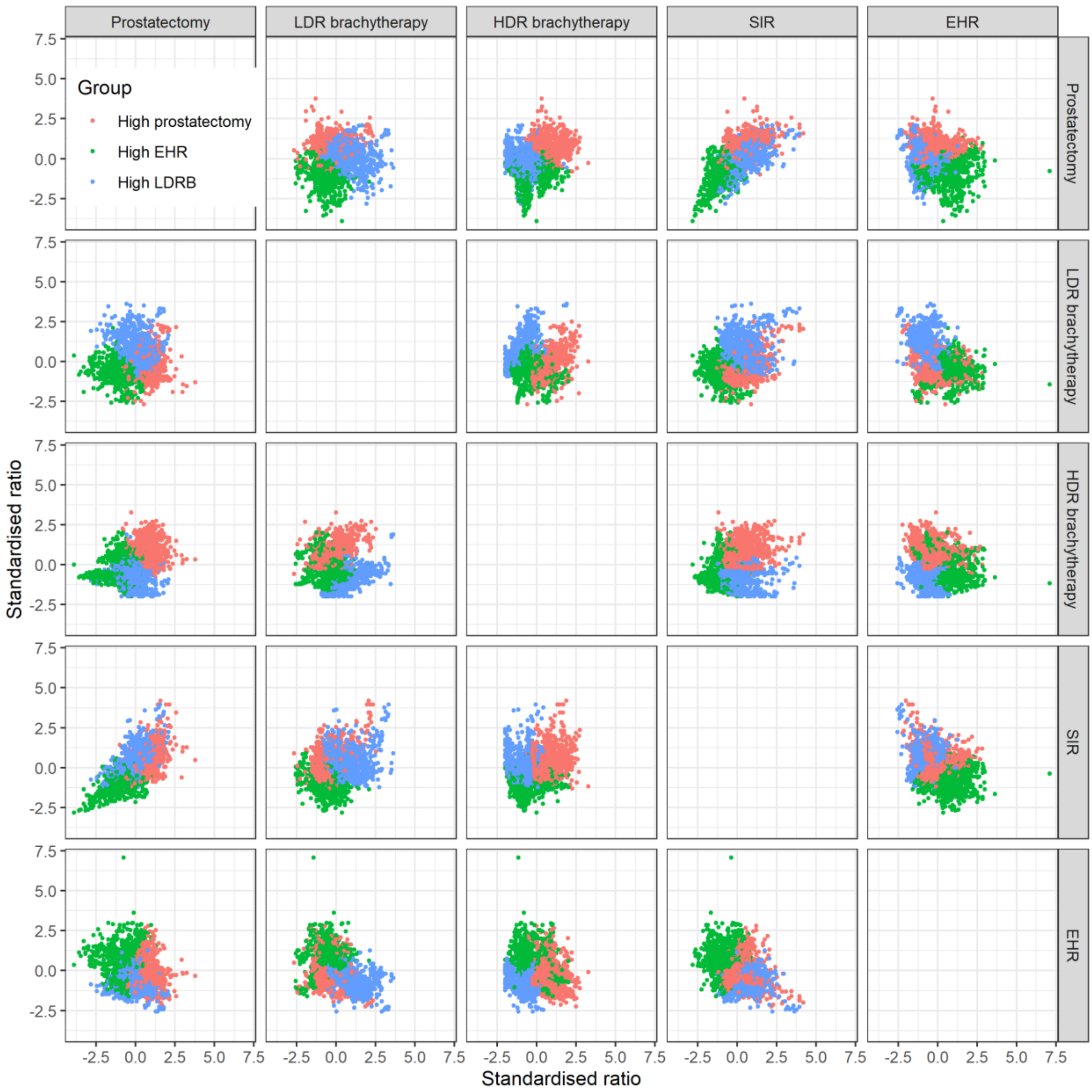


*Figure S1; Pairwise scatterplots of area-level SSRs for the treatments for prostate cancer and standardised incidence ratios (SIRs) for prostate cancer diagnoses and excess hazard ratios (EHRs), with each dot in each subplot reflecting a single area and the symbol colour reflecting the area's statistical group. Note that the standardised ratios shown here have been normalised and log transformed prior to normalisation.*

### Methods

#### Medical coding

*Table S2; ICD-10-AM/ACHI diagnosis related group codes for treatments for prostate cancer.*

| Treatment | ICD-10-AM/ACHI codes |
|---|---|
| High dose rate brachytherapy | 15327-01, 15327-03, 15327-06, 15327-07, 15536-00, 15536-01, 15536-02 |
| Low dose rate brachytherapy | 15327-00, 15327-02, 15327-04, 15327-05, 15338-00 |
| Radical prostatectomy | 37209-00, 37210-00, 37211-00, 37209-01, 37210-01, 37211-01 |

#### Areal reallocation

This section describes in greater detail the reallocation from SLA to 2011 ASGS SA2. Let's describe the SLA as the "original area" ($A_{Oi}$) and the 2011 ASGS SA2s as the "final area" ($A_{Fi}$).

If area $A_{Oi}$ overlaps $\boldsymbol{A_{Fi}} = \{A_{Fi1}, A_{Fi2}, \dots, A_{FiM}\}$, a set of $M$ SA2s from the 2011 ASGS and the proportion of all individuals living in $A_{Oi}$ who live in each area of $\boldsymbol{A_{Fi}}$ is

$$\boldsymbol{P_i} = \{P_{i1}, P_{i2}, \dots, P_{iM}\}$$

where $\sum_{m=1}^{M} P_{im} = 1$

Then we define the cumulative proportions as:

$$P'_{\text{im}} = \sum_{v=1}^{m} P_{iv} \quad for\ all\ m\ in\ 1, \dots, M$$

Additionally, $P'_{i0} = 0$.

For each individual in the data set, a random number, $rand$, is generated using a uniform distribution between 0 and 1. An individual who was originally geocoded to $A_{Oi}$ is then reallocated to $A_{Fim}$, such that $P'_{\text{im}} \geq rand$ and $P'_{i(\text{m}-1)} < rand$

#### Spatial modelling

For each of the procedures, the expected number of separations ($E_i$) was calculated for each SA2 ($i$, 2011 ASGS) for the ten-year period between 2007 and 2016, using the Australian total count ($y_k$) and population ($pop_k$) for each age group ($k$) and the population in the SA2 at each age group ($pop_{ik}$) as follows.

$$E_i = \sum_k \frac{y_k}{pop_k} pop_{ik}$$

*Equation 1*

The observed total number of separations in each SA2 ($y_i$) was modelled as a Poisson process with an intensity proportional to $E_i$.

$$y_i \sim Poisson(E_i \theta_i)$$

*Equation 2*

The log of the standardised separation rate ratio ($\theta_i$) was a linear combination of an intercept ($\beta_0$) and a spatial random effect ($S_i$).

$$\log(\theta_i) = \beta_0 + S_i$$

*Equation 3*

A Leroux prior was applied to $S_i$, so that the spatial effect for area $i$ depended on the spatial effects for all other areas ($S_{\setminus i}$), according to a neighbourhood matrix. The neighbourhood matrix was square with dimension equal to the number of SA2s being modelled. The $(i, j)^{\text{th}}$ element of the matrix ($w_{ij}$) was 1 if $i^{\text{th}}$ and $j^{\text{th}}$ SA2s were adjacent and 0 otherwise. The parameter $\rho$ reflected the degree of spatial autocorrelation and $\sigma_S^2$ was the variance in the spatial random effects.

$$S_i | S_{\setminus i} \sim \mathcal{N}\left(\frac{\rho \sum_j w_{ij} S_j}{\rho \sum_j w_{ij} + 1 - \rho}, \frac{\sigma_S^2}{\rho \sum_j w_{ij} + 1 - \rho}\right)$$

*Equation 4*

Weakly informative priors were applied, as follows, where $\mathcal{IG}$ is the inverse gamma distribution and $\tau_S^2$ was the precision in the spatial random effect.

$$\beta_0 \sim \mathcal{N}(0, 100\,000)$$

$$1/\sigma_S^2 = \tau_S^2 \sim \mathcal{IG}(1, 0.01)$$

$$\rho \sim Uniform(0, 1)$$

*Equation 5*

#### Convergence

Within chain convergence of the Markov chain Monte Carlo was confirmed using the Geweke diagnostic, with less than 10% of areas' having a significant Geweke diagnostic at the 5% level. Trace plots and posterior densities of the global parameters ($\beta_0$, $\rho$ and $\sigma_S$) and a small number of area-specific parameters ($\theta_i$ and $S_i$) were visually inspected.

#### Sensitivity analysis

A sensitivity analysis was conducted to test whether the model was sensitive to choices of the parameter values for the priors on $\beta_0$ and $\tau_S^2$ in Equation 5. The values trialled are shown in Table S3 and the results are shown in Figure S2, Figure S3 and Figure S4. Sensitivity analyses showed that the results were not sensitive to the choice of priors or hyperpriors. Similarly, the results were not sensitive to initial values or seed.

*Table S3; Values for the hyperpriors trialled in the sensitivity analysis.*

| | Mean | Variance |
|---|---|---|
| $\beta_0$ | -1; 0; 1; 2 | 50; 1000; 10 000; 100 000 |
| | Scale | Shape |
| $\tau_S^2$ | 0.00001; 0.001; 0.01; 0.1 | 1; 2; 3; 4 |

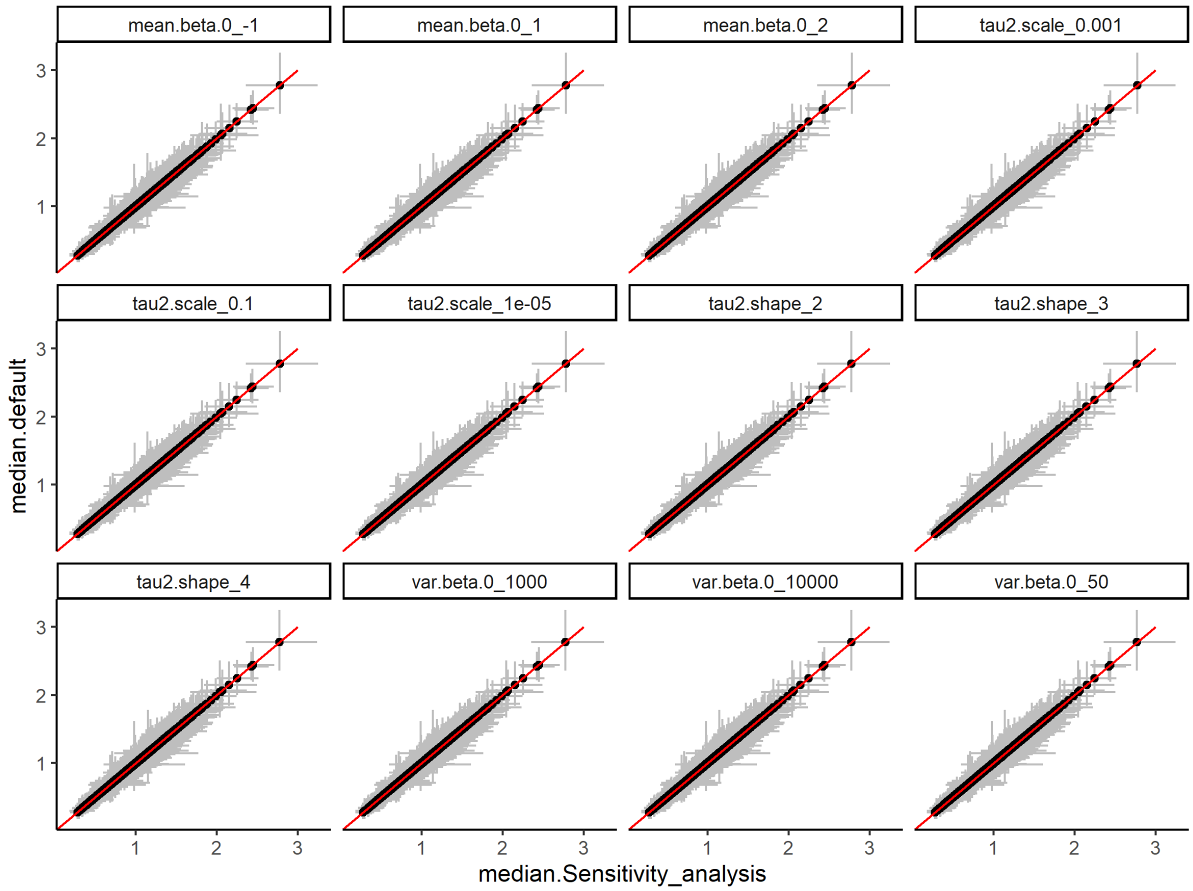


*Figure S2; Results of the sensitivity analysis for radical prostatectomy. The y-axis shows the median SSR (and 95% credible interval) for each area using the default value for the hyperprior and the x-axis shows the median SSR (and 95% credible interval) for each area using the hyperprior value specified in the label for the subplot. The line of equality is shown in red to guide the eye.*

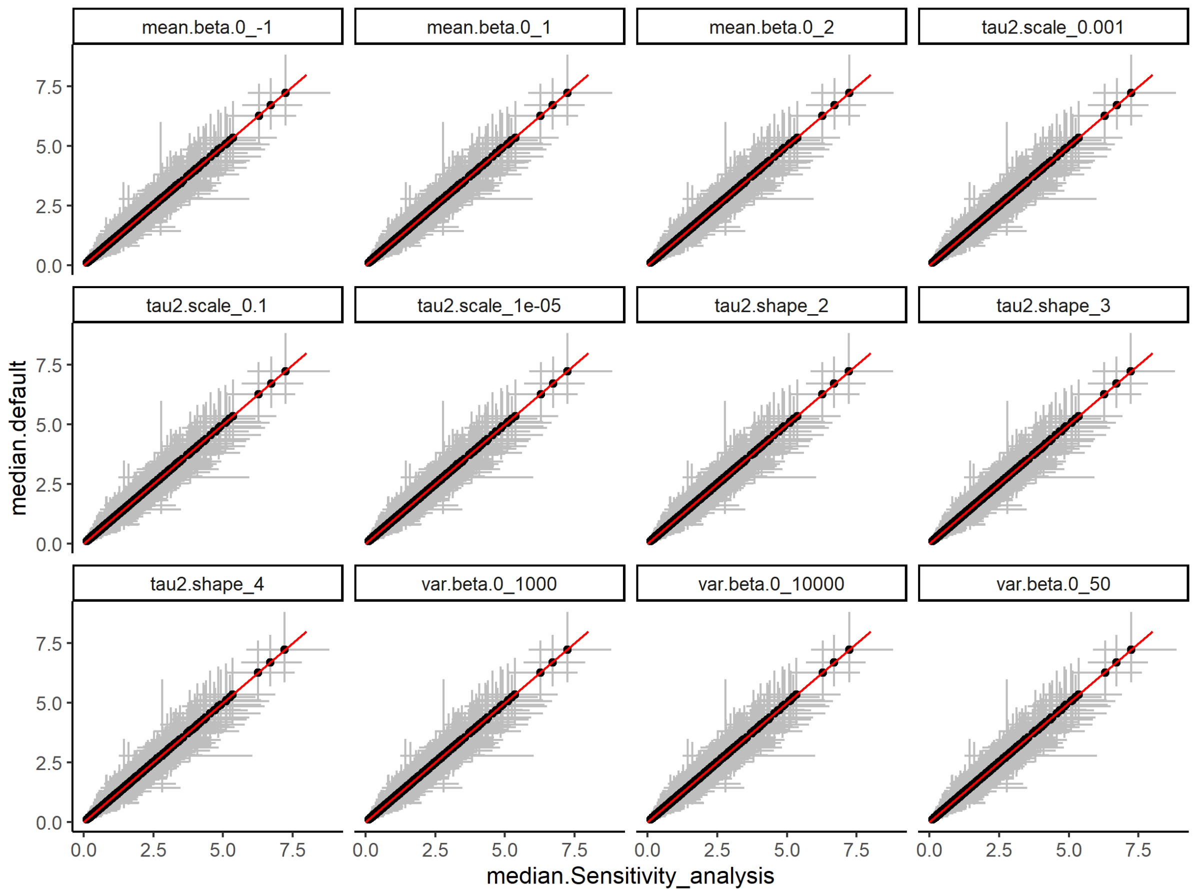


*Figure S3; Results of the sensitivity analysis for low dose rate brachytherapy. The y-axis shows the median SSR (and 95% credible interval) for each area using the default value for the hyperprior and the x-axis shows the median SSR (and 95% credible interval) for each area using the hyperprior value specified in the label for the subplot. The line of equality is shown in red to guide the eye.*

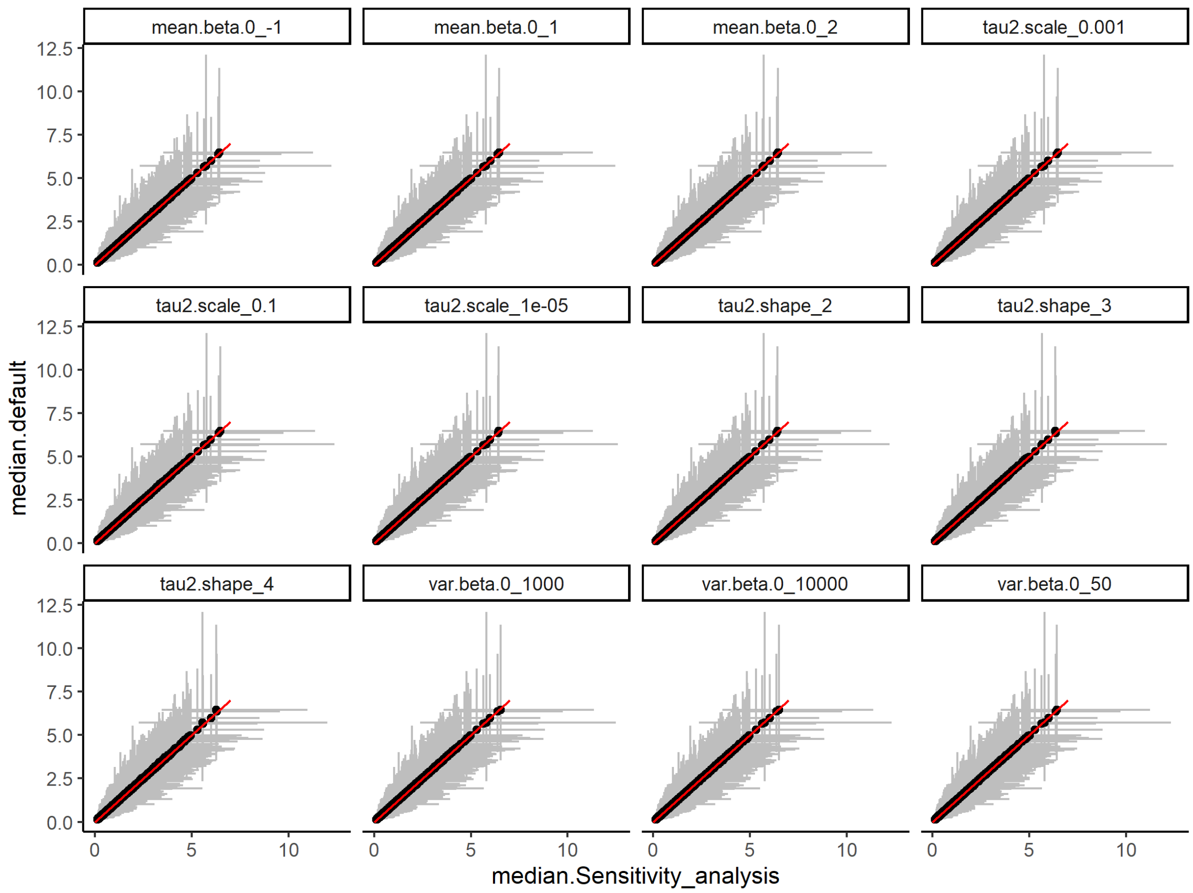


*Figure S4; Results of the sensitivity analysis for high dose rate brachytherapy. The y-axis shows the median SSR (and 95% credible interval) for each area using the default value for the hyperprior and the x-axis shows the median SSR (and 95% credible interval) for each area using the hyperprior value specified in the label for the subplot. The line of equality is shown in red to guide the eye.*
